# Assessing Senegal’s Performance Against the 7-1-7 Benchmark for Outbreak Preparedness: A Retrospective Analysis Using National Surveillance Data (2022–2023)

**DOI:** 10.1101/2025.04.10.25325599

**Authors:** Jerlie Loko Roka, Yoro Sall, Youssou Bamar Gueye, Khadim Kebe, Diop Boly, Aida Kanoute, Ibra Diagne, Papa Samba Dieye

## Abstract

This study assesses Senegal’s performance against the 7-1-7 approach, a benchmark for outbreak preparedness, focusing on infectious disease detection, notification, and response from 2022 to 2023. Using secondary national surveillance data, the analysis included socio-demographic, clinical, and laboratory information, with data visualized using ArcGIS.

Findings showed a median detection period of 12 days, twice the recommended duration, due to factors like healthcare-seeking behavior and lab processing times. Notification was relatively efficient, with a median of 1 day post-lab confirmation. However, delays in activating the national Emergency Operations Center (EOC) were noted.

Despite limitations such as incomplete arbovirus data and an unstructured response data system, the study highlights the need to align current surveillance indicators with the 7-1-7 metrics, digitalize the notification system, and empower additional laboratories. These interventions are essential for improving public health responses, enhancing health outcomes, and increasing community resilience in Senegal.

## Introduction

A global effort, involving more than 70 countries worldwide, was initiated through the Global Health Security Agenda (GHSA) framework in 2014. The aim was to strengthen countries’ capacity to prevent, detect, and respond to public health threats, with a specific focus on infectious diseases, contributing to a safer world ^1^. The detection component includes diseases surveillance which is key for public health decision making and is defined by the World Health Organization (WHO) as ‘the ongoing systematic identification, collection, collation, analysis and interpretation of disease occurrence and public health event data, for the purposes of taking timely and robust action, such as disseminating the resulting information to the relevant people, for effective and appropriate action’ ^2^. The foundation of effective public health surveillance lies in the recognition of valid information as a cornerstone for timely decision-making. In the realm of public health, the ability to gather, analyze, and interpret accurate data is vital for identifying potential threats, understanding emerging trends, and implementing prompt and targeted interventions. This emphasis on valid information not only ensures the reliability of surveillance systems but also plays a crucial role in safeguarding community health by facilitating informed and strategic decision-making processes ^3^. Timeliness is a quantitative attribute evaluated in public health surveillance assessments. However, it is notable that there is currently no universally standardized method for assessing timeliness in the context of public health surveillance. Additionally, the timeliness of reporting may vary from one disease to another, underscoring the necessity for disease-specific analyses to better tailor recommendations ^4 5^. The 7-1-7 approach has been proposed as a standard time benchmark for assessing a country’s preparedness in managing suspected outbreaks. According to this metric, a suspected outbreak should be detected within 7 days of emergence, reported to public health authorities within 1 day of detection, and seven early response actions should be completed within 7 days from reporting to public health authorities ^6^. The detection of suspected outbreaks is gauged by the time elapsed from exposure to the disease agent to the initiation of a public health intervention. This process comprises several key milestones, including onset of symptoms, onset of behavior, capture of data, completion of data processing, capture of data in the surveillance system, application of pattern recognition tools/algorithms, and generation of automated alerts ^7^. An evaluation of the 7-1-7 approach has been conducted to assess feasibility in five countries. The findings highlight that vaccine-preventable diseases and vector-borne diseases surpassed the 7-day target and may be of interest in subsequent analysis ^8^.

This specific analysis has never been conducted in Senegal where the public health surveillance system was established back in 1979, initially focusing on vaccine-preventable diseases. The primary objective of the current study is to document the timeline of detection, notification, and response for four diseases in Senegal. Additionally, the study aims to conduct an analysis to identify weaknesses and provide guidance to enhance the country’s readiness.

## Methodology

### Study setting

Since it was established, the surveillance system has evolved and is now aligned with the World Health Organization’s (WHO) Integrated Disease Surveillance and Response (IDSR) framework. Currently, the surveillance system encompasses 52 diseases, with 22 of them being reported on a weekly basis. Both passive and active surveillance strategies are applied in the country with the active surveillance being conducted mostly during the sites visit that are identified by each district and follow the acute flaccid paralysis (AFP) sites as a driver. While the indicator based was well established its extension to the community level started only in 2016 using the health community workers. In addition to the indicator-based surveillance (IBS), an event-based surveillance (IBS) set up and implementation started during the COVID-19 pandemic that accelerated its process. However, in post pandemic period, the functionality is limited, and analysis is ongoing to identify bottle necks and way forward. A sentinel syndromic surveillance system is in place and had a crucial role in the country for public health threats detection. The country currently had 26 sentinel’s sites in all regions and had developed a scaled-up plan for the 5 coming years to ensure the best representativity of the country.

The Emergency Operations Center (EOC) has been officially established in 2014 at national level and was activated 2 times since then. Five decentralized, subnational EOCs were established start from 2021 with the target of having an EOC in each of the 14 regions. Each decentralized EOC activation is under the regional governor responsibility and is done with the technical support of the national EOC that provided guidance and tools when required.

### Case definition and surveillance system information flux

For each of the diseases monitored through the formal Integrated Disease Surveillance (IBS), including the sentinel surveillance, a precise case definition has been developed in accordance with the World Health Organization (WHO) generic definition and guidance and may evolve over time. The country has established case definitions tailored for the community level; alongside suspected case definitions utilized within health facilities. However, it’s noteworthy that only 8 diseases/events are currently under surveillance at the community level while all the 52 are concerned at health structures level. Whenever a case aligns with the community case definition, it is promptly referred to the nearest health facility, where a healthcare professional assesses the case. Upon detection of a suspected case, the requisite sample collection is carried out and dispatched to the reference laboratory for confirmation. In Senegal, the Institut Pasteur de Dakar (IPD) serves as the reference laboratory for all viral diseases under surveillance. Positive results are communicated to the national-level team responsible for disease surveillance (the Directorate of Prevention within the Ministry of Health), which subsequently shares the information with the respective region and district medical chiefs for necessary action, including preliminary investigation. Based on the reference technical group recommendations^7^, we considered as detection the timeline from the disease onset to the laboratory confirmation without which detection is not possible. We considered the communication of those laboratory results to the national level health authorities as the notification and finally a formal incident management system (IMS) activation has been taken into consideration. However, as the country still working on its IMS network and has not fully integrated the use of the IMS for all public health threats, we took into consideration responses that may have been set and conducted without a formal IMS.

### Study period and data collection

This study analyzes public health threat events in Senegal during 2022 and 2023, utilizing secondary analysis of national case-based surveillance data for various diseases. The data, extracted from distinct databases for each disease, were de-identified and shared for the study. Each dataset includes socio-demographic and clinical data, such as the date of onset, laboratory results dates, and information sharing with public health authorities. Due to missing key dates in early 2022, complete data for Hemorrhagic Viral Fever (HVF) were considered starting from July 2022. Adverse Events Following Immunization (AEFIs) were excluded from the analysis as their detection is purely clinical and does not require laboratory confirmation. Additionally, all AEFIs identified during the study period were COVID-19 related, which may not reflect typical, routine situations.

The study extracted dates for sample reception at laboratories and when results became available, as well as dates when laboratory results for positive cases were notified to the Ministry of Health via email communications. A desk review of the Incident Management System (IMS) report notes was conducted to gather information on national Emergency Operations Center (EOC) activations. Information on responses conducted without an IMS was also collected to ensure all actions were accurately captured. The collated data were compiled into a new Excel sheet, enabling an assessment of response timelines for various events.

### Data analysis

To enhance the analysis of the detection timeframe, we divided the timeline into three distinct categories. First, we calculated the timeline between the date of onset and the contact with health facilities. Subsequently, we computed the timeline between the onset and sample collection, and finally, the timeline between the onset and the availability of laboratory results. Regrettably, we were unable to assess the timeline between the onset and the date the laboratory received the samples, as this data was not captured in the dataset that was made available for this study. The median duration in days for each of these timelines is presented as a result. Furthermore, all our analyses were conducted exclusively on laboratory-confirmed cases. We consolidated all the diverse datasets into a single dataset, ensuring that identical columns were merged seamlessly and accurately. Analyses were performed using python. Additionally, we visually represented the detection performance of each district using ArcGIS, focusing on the timeline between the onset and the laboratory result date. This was achieved through a density map, where districts achieving the target of 7 days for detection were colored in green, those falling between 8 and 14 days were colored in orange, and districts exceeding 14 days for detection were colored in red. This mapping exercise provides a clear and informative visualization of the efficiency in disease detection across different districts.

### Ethical

The Senegalese Ministry of Health, Department of Prevention, granted permission to use deidentified case base surveillance data collected during the study period in the country. No local ethical committee approval was required for this secondary data analysis of routine surveillance data.

## Results

### Detection and notification

Our study identified five major public health events during the study period. It’s important to note that measles occurred repeatedly in the same location and more frequently in different locations. Our analyses showed that in median a patient seeks for care in a health facility 3 days after the disease onset except for yellow fever (11 days) and Crimea-Congo (6 days). The longest detection period was observed for Crimea-Congo fever, with a median of 27 days. In contrast, Chikungunya had the shortest detection period, typically identified within 10 days. While the duration from patient presentation at a health facility to detection is significant across all infectious public health events, it’s noteworthy that yellow fever exhibited the shortest delay, with laboratory results available within just 2 days of seeking care. Meanwhile, this period extended to 21 days for Crimea-Congo fever. During the study period, the country recorded 1022 cases of measles IgM positivity. Patients with measles sought care within a median of 3 days; however, it took a median of 10 days from onset to availability of laboratory results (table 1). Our analysis revealed that only one district met the criteria for detecting public health events within 7 days from the disease onset to the availability of laboratory results at the laboratory level. Fifty-six districts detected events within 2 weeks (14 days). No public health events were detected in 15 health districts during the study period. A total of 16 districts detected public health events more than 2 weeks after the disease onset. Only a few (five districts) detected public health events within 7 days. The concerned districts are in the Dakar region (Mbao, Sangalkam, and Dakar Ouest), Louga region (district of Louga), and Sedhiou region (District of Goudomp) (Figure 1).

**Table 1:** Public health events metrics for detection and notification, Senegal, 2022-2023.

| Public health event | Onset to HF visit<br>Median days(range) | Onset to lab results<br>Median days (range)<br>Target 7 days | detection to notification Median<br>days (range)<br>Target 1 day |
| --- | --- | --- | --- |
| Measles (n=1022) | 3 (0-65) | 10 (0-72) | 1(0-20) |
| Yellow fever (n=3) | 3 (1-5) | 11 (9-27) | 0 |
| Dengue (n=319) | 2 (0-22) | 12 (1-74) | 0 |
| Chikungunya (n=288) | 1 (0-24) | 11 (1-40) | 0 |
| Crimea-Congo (n=11) | 4 (1-10) | 11 (6-21) | 0 |
| Rift Valley Fever (n=3) | 3(2-4) | 10(8-13) | 0 |

**Figure 1.**
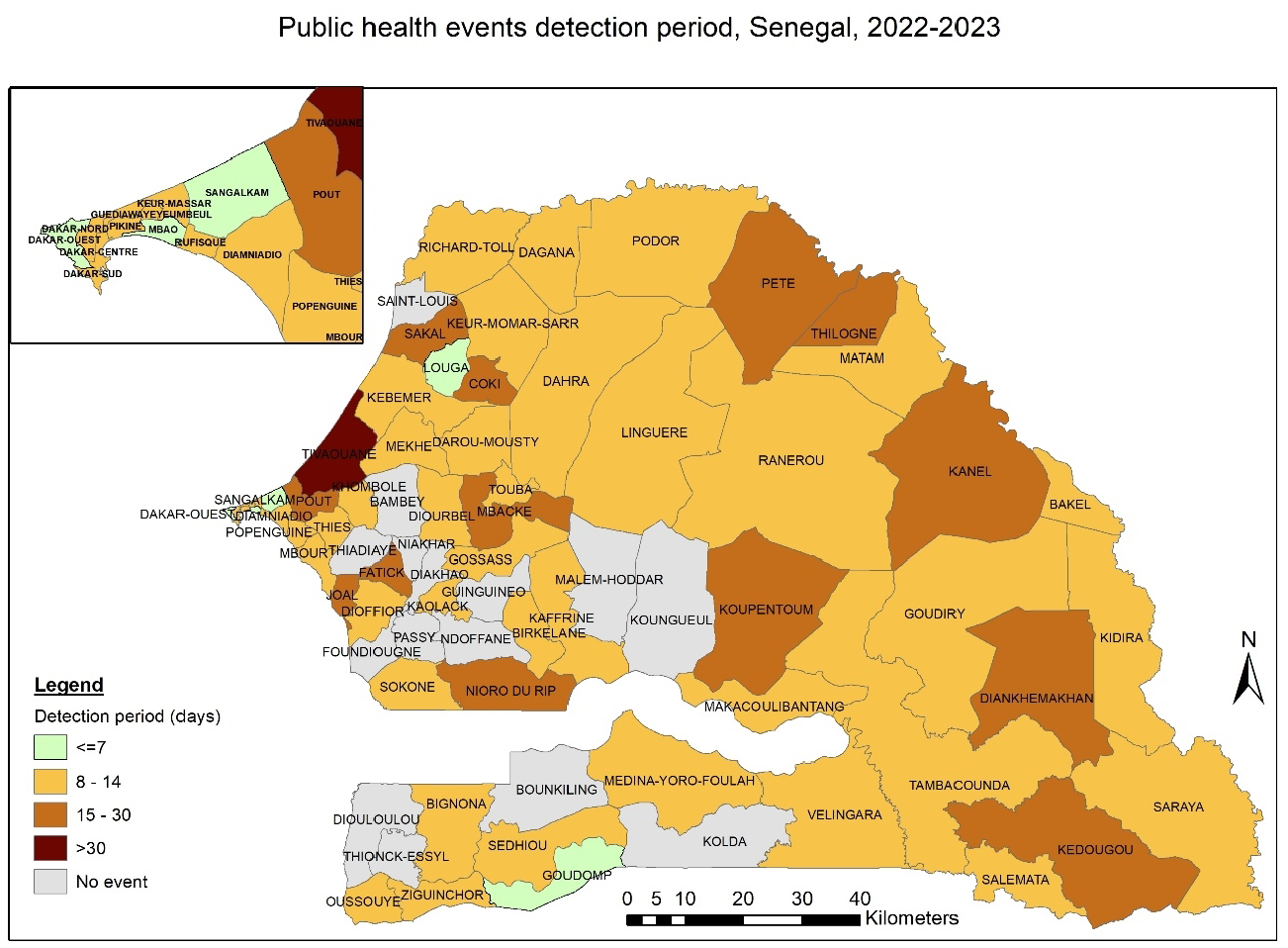
Mapping of Public Health detection period at the district Level: Senegal, 2022-2023.

Notification was generally done immediately after the laboratory results were available. However, for measles, notification varied from the same day to a maximum of 20 days. Apart from measles, notification was done as soon as results were available (table 1).

### Response

Throughout the study period, sub-national Emergency Operations Centers (EOCs), implemented starting in 2021, were activated three times: twice in response to the avian flu H5N1 outbreak detected in wildlife (that is not included in the current paper) and once for Crimea Congo (table 2). At the subnational level, activation occurred in 2 days after laboratory results were issued.

**Table 2:** Timeline of National and Sub-National Activation and Initial Public Health Actions.

| Event | Activation Level | Notification to Activation (days) | Notification to first PH actions (days) |
| --- | --- | --- | --- |
| Yellow fever | No activation |  | Selective response |
| Measles | No activation |  | Local response |
| Rift Valley fever | No activation |  |  |
| Chikungunya | No activation |  |  |
| Crimea Congo | National EOC | 7 | 5 |

The national PHEOC was activated once during the study period and twice since its implementation. Activation occurred 7 days after the laboratory results were issued and 6 days after the preliminary rapid case assessment. The full multidisciplinary case investigation was still ongoing when the EOC was activated and lasted for a week. Initial actions, including contact tracing, secured burial measures, and reinforcement of Infection, Prevention and Control (IPC) measures, began during the preliminary rapid case assessment. No risk assessment was included during the preliminary rapid case assessment.

Concerning measles and other vaccine-preventable diseases, local investigations followed by reactive vaccination campaigns were carried out despite the absence of a formal IMS.

For all events including measles outbreak, an investigation was conducted or by a local team or both local, regional and national team when required. Each investigation ended with a risk assessment using the WHO STAR tool^1^.

## Discussion

### Detection

In Senegal, the detection of public health events, regardless of their type, typically occurs within a median period of 12 days(min:7-max:49), which is almost twice the expected time based on the 7-1-7 metrics. A detailed analysis can only be conducted by breaking down and understanding the median period spent by patients in their journey from disease onset to detection by the health system.

Healthcare-seeking behavior represents the initial crucial step in facilitating disease detection, even if community health workers fulfill their roles of identifying and directing patients to health facilities. The overall period of health care seeking in Senegal is acceptable (around 3 days with a maximum of 6 days), however the delay registered for some of the disease highlight the need to pursue efforts in improving health facilities frequentation. Health care seeking behavior can be influenced by diverse factors that vary from one region to another. Studies conducted in the West and Central African regions have revealed a generally low percentage of healthcare-seeking behavior with less than 40% of patients seeking for health in Senegal for fever^9-11^. Furthermore, factors including distance, maternal education level, household wealth status, and parental age have been identified as significant determinants of healthcare-seeking behavior in the region ^9^. An overarching development program extending beyond the confines of the health sector and encompassing the construction of health facilities and provision of equipment closer to the population, as well as educational initiatives targeting girls are necessary to mitigate delays. Meanwhile, community health workers can play a pivotal role in identifying symptomatic patients and directing them to health facilities, while concurrently engaging in health promotion communication within the community as shown in different studies ^12 13^.

Our analysis indicates that the required samples were mostly collected within 1 day starting from the consultation day for all public health events included in our study, except for Crimea-Congo. In the case of Crimea-Congo, samples were collected 4 days after the consultation day. This observation may be attributed to difficulties faced by healthcare personnel in identifying the patient as a suspected case that needs to be reported. The country has developed case definitions for all diseases under surveillance, personal have been trained at all levels of the health pyramid and supervisions are conducted. However, the country is still experiencing delays, particularly in hospitals. Training and involving first-line physicians in public health disease surveillance and detection remain challenging as reported in similar by other studies ^14-16^. Indeed, physicians primarily focus on treating and curing their patients. Therefore, understanding infectious disease notification or surveillance systems becomes less important, and in many hospitals, it is not conducted or considered a priority. The Senegalese surveillance division is not only working to include physicians in surveillance training sessions, but, more importantly, is advocating to raise awareness among them. Additionally, efforts are being made towards the identification of surveillance focal points in each hospital.

Except for the Crimean-Congo case, laboratory results were issued within a period ranging from 2 to 10 days from the day the patient specimen was collected for all other public health events. According to WHO guidelines, when specified, surveillance performance indicators are set for laboratory results (IgM or RTPCR) to be issued within 4 days of receipt of specimen for yellow fever or measles. The same documents are targeting a period of 3 to 5 days for specimens to be received at the laboratory after collection ^17 18^. The current WHO surveillance performance indicators guidance renders detection within 7 days inconsistent, particularly given that the median period for seeking care in our context is 3 days, and laboratory results are rarely available within one day of specimen reception. This underscores the necessity to harmonize and revise current surveillance performance guidance to assist countries in aligning with and achieving the 7-1-7 metrics.

### Notification

Our analysis showed the median notification period of 1 day after the result is available at the laboratory is satisfactory and aligned with the 7-1-7 metrics. However, the wide range suggests room for improvement. Many factors may have played a role, from reagent stockouts to the potential increase in the number of samples processed by the only reference laboratory in the country, specifically in the case of measles. Empowering other laboratories could be an option for reducing delays and improving notification metrics. In addition, the current notification system relies solely on email sharing among stakeholders, whereas a digitalized platform with automatic messages to key stakeholders in the event of a positive laboratory result may provide a better-structured approach, minimizing potential delays in the notification process. The importance of a structured system becomes evident when the country faces a significant number of positive cases to report, as demonstrated by the measles laboratory results, which had the widest range of notifications.

### Response

The national-level EOC, implemented in December 2014 as a public health emergency response tool during the Ebola outbreak in West Africa, has been activated twice over its decade of functionality: once for COVID-19 and once for Crimean-Congo hemorrhagic fever. National PHEOC activation in Senegal is less frequent compared to countries like Cameroon and Uganda, which implemented their PHEOC around the same period as Senegal but have experienced multiple activations. In 2016 alone, the Cameroon PHEOC was activated 9 times, while Uganda’s PHEOC was activated for 271 public health events between 2014 and 2021^19,20^. Many reasons can explain the infrequent activations in Senegal, including a lack of understanding and appropriation of the PHEOC or existing bodies like the national committee for outbreak management. The existence of this latter body can lead to confusion regarding the national EOC’s functionality, despite both bodies having well-defined missions on paper. It is up to the country’s political and health authorities to decide on the best way to utilize this tool and provide appropriate guidance.

During our study period, the national EOC was activated once, in response to the Crimean-Congo hemorrhagic fever outbreak. It was activated 7 days after laboratory results and 5 days after the case rapid assessment later after the recommended timeline of activating a public health Emergency Operation Centers within 120 minutes^21^. Despite many countries being described as having fully functional Public Health Emergency Operation Centers (PHEOCs) ^22^, we found very few that have documented monitoring of their performance regarding the 7-1-7 metrics, and more specifically, the PHEOC activation timeline. However, Cameroon has managed to significantly reduce its activation timeline from 8 weeks to 24 hours within one year ^20^. Despite the lack of formal documentation, the experiences of Uganda and Cameroon highlight the critical role of staff training, multiple simulations, frequent activation of the PHEOC, and strong coordination in the PHEOC implementation process and its success in better supporting emergency management ^19^. In addition, delays experienced in national PHEOC activation can be shortened by including a rapid risk assessment in the preliminary investigation. This would guide fast decision-making instead of waiting for a multidisciplinary, full-scale investigation that could last many days.

## Limitations

The study encountered significant challenges due to the lack of systematic data collection for arboviruses and public health notifications, necessitating reliance on emails for tracking. Additionally, there was no structured system for documenting response timelines and actions. Only human-related public health events were included, excluding One Health (OH) events like the H5N1 outbreak, which require different response mechanisms and could bias the analysis. These limitations underscore the need for improved systematic data collection and integrated response strategies to enhance public health surveillance and response accuracy in Senegal.

## Conclusion

Our study evaluates Senegal’s public health system, emphasizing detection, notification, and response to infectious diseases. Findings reveal significant delays in event detection, with a median period of 12 days from disease onset, double the duration recommended by the 7-1-7 metrics. Factors contributing to this delay include healthcare-seeking behavior and challenges in sample collection and laboratory processing. Notification practices were relatively efficient, with a median period of 1 day post-laboratory confirmation, though further improvement is needed. Response capabilities, especially the national Emergency Operations Center (EOC) activation, showed delays, highlighting the need for rapid response improvements. Comparisons with countries like Cameroon and Uganda emphasize the importance of regular training and coordinated efforts for better EOC activation and emergency management. Despite limitations, this study provides insights into the strengths and weaknesses of Senegal’s public health system, advocating for targeted interventions to enhance effectiveness and improve readiness and response to public health threats.

## Data Availability

All data and related metadata underlying the findings reported in this manuscript will be available upon request

## Footnotes

1 https://www.who.int/publications/i/item/9789240036086

